# Inconsistency detection in cancer data classification using explainable-AI

**DOI:** 10.1101/2024.10.02.24314783

**Authors:** Pouria Mortezaagha, Arya Rahgozar

## Abstract

This paper presents a novel approach to improving text-based cancer data classification by integrating BERTopic clustering with Support Vector Machine (SVM) classifiers, combined with the Explainable Inconsistency Algorithm (EIA). The proposed method leverages advanced preprocessing techniques, including Node2Vec embeddings, to enhance both clustering and classification performance. Through the introduction of EIA, we automatically identify and eliminate outliers and discordant data points, thus improving classification accuracy and providing valuable insights into underlying data relationships. A key innovation in this work is the use of recommender systems for mapping clusters to labels, which improves label assignment through collaborative filtering techniques. Our experimental results show a significant increase in both accuracy and F1-score after addressing data inconsistencies, with improvements validated through statistical tests, including t-tests. This paper contributes a robust, explainable, and scalable framework for cancer data analysis, offering potential applications in other domains requiring high-precision text classification. Future work will focus on extending the EIA to other biomedical datasets, optimizing hyperparameters, and deploying the framework in real-time clinical decision-support systems.

## 1 Introduction

In machine learning and data science, integrating multiple approaches can overcome the inherent limitations of any single method, leveraging the strengths of each to improve overall performance. This paper proposes a dual-perspective model to address classification challenges by decoupling distinct sets of features and employing different algorithms: one with labeled data (supervised learning) and the other without labels (unsupervised clustering). By using clustering techniques, we aim to detect, minimize, and mitigate bias, outliers, noise, or inconsistencies, all of which are commonly embedded within the labels [Hellström et al., 2020, He et al., 2002, Wang et al., 2019, Naseem et al., 2023a, Feldmann et al., 2015]. In this context, we use these terms interchangeably to describe discrepancies in taxonomic perspectives, whether in statistics, machine learning, or requirements engineering.

This research integrates a Large Language Model (LLM), specifically BERT, with a traditional machine learning algorithm to introduce a new dimension of analysis. We have chosen Support Vector Machines (SVM) as our traditional machine learning model, which is widely recognized for its effectiveness in high-dimensional spaces and its versatility in handling various data types through the selection of kernel functions [Cervantes et al., 2020]. Despite its robustness, SVM faces challenges with scalability and kernel selection, particularly when working with massive datasets, which can result in prohibitive training times and complexity in preprocessing [Wen, 2023].

To complement SVM, we employ BERT (Bidirectional Encoder Representations from Transformers), which has revolutionized natural language processing by offering a bidirectional context analysis, allowing it to understand the meaning of words based on their surrounding text [Devlin et al., 2019]. This makes BERT especially effective at capturing subtle semantic relationships in the data [González-Carvajal and Garrido-Merchán, 2023]. However, LLMs like BERT also present challenges, such as susceptibility to overfitting and the potential for suboptimal generalization when fine-tuning using cross-entropy loss [Liu et al., 2017, Naseem et al., 2023a].

To address these challenges, we employ BERTopic [Grootendorst, 2022] for clustering the semantic-rich embeddings produced by BERT. BERTopic distinguishes itself as a sophisticated topic modeling technique that combines clustering with a class-based variation of TF-IDF to generate coherent and meaningful topic representations [Samsir et al., 2023]. By incorporating BERTopic, we not only add a new perspective to the traditional machine learning approach but also provide interpretability by linking clusters to associated terms, which helps explain the inconsistencies between the two models.

In addition to BERT’s semantic strengths, we utilize knowledge graphs and Node2Vec embeddings [Grover and Leskovec, 2016] to enhance feature engineering for SVM. Knowledge graphs and Node2Vec provide a structural and relational view of the data, capturing connections between entities, while also offering scalability and robustness for tasks requiring graph-based learning [Palumbo et al., 2018, Wang et al., 2021]. This dual perspective allows us to leverage both linguistic and structural insights for more comprehensive data analysis.

By integrating SVM’s mathematical precision with the relational insights from knowledge graphs and the linguistic depth of BERT, we create a twin machine learning pipeline that captures different viewpoints on the classification of data instances. Instances where the models diverge in their classifications are labeled as “inconsistent,” and we hypothesize that eliminating these inconsistencies will lead to improved predictive performance, which will be reflected in our evaluation metrics.

### 1.1 Objectives

- Automatic multi-perspective inconsistency detection and classification refinement through the design of a dual-algorithmic pipeline, combining classification with clustering.
- Provide explanations for predictions using LLM-based clustering techniques.
- Ensure flexibility to incorporate any machine learning model or LLM into the framework.
- Feature refinement to improve classification performance.

A key feature of our methodology is the strategic removal of inconsistent data, followed by a comprehensive evaluation of its impact on performance metrics. Figure 1 presents the overall system architecture. The process begins with dataset preprocessing, including subject-verb-object (SVO) extraction and knowledge graph construction using tools like SpaCy^1^, NetworkX^2^, and Pyviz^3^. Node2Vec embeddings [Grover and Leskovec, 2016] are then generated, feeding into a classification model. Simultaneously, BERTopic clustering is applied, with the resulting clusters mapped to human annotations. This cluster-label mapping is incorporated into a recommender system using integer programming, allowing for the identification and removal of inconsistent nodes, ultimately leading to refined classification results validated through statistical testing.

**Figure 1.**
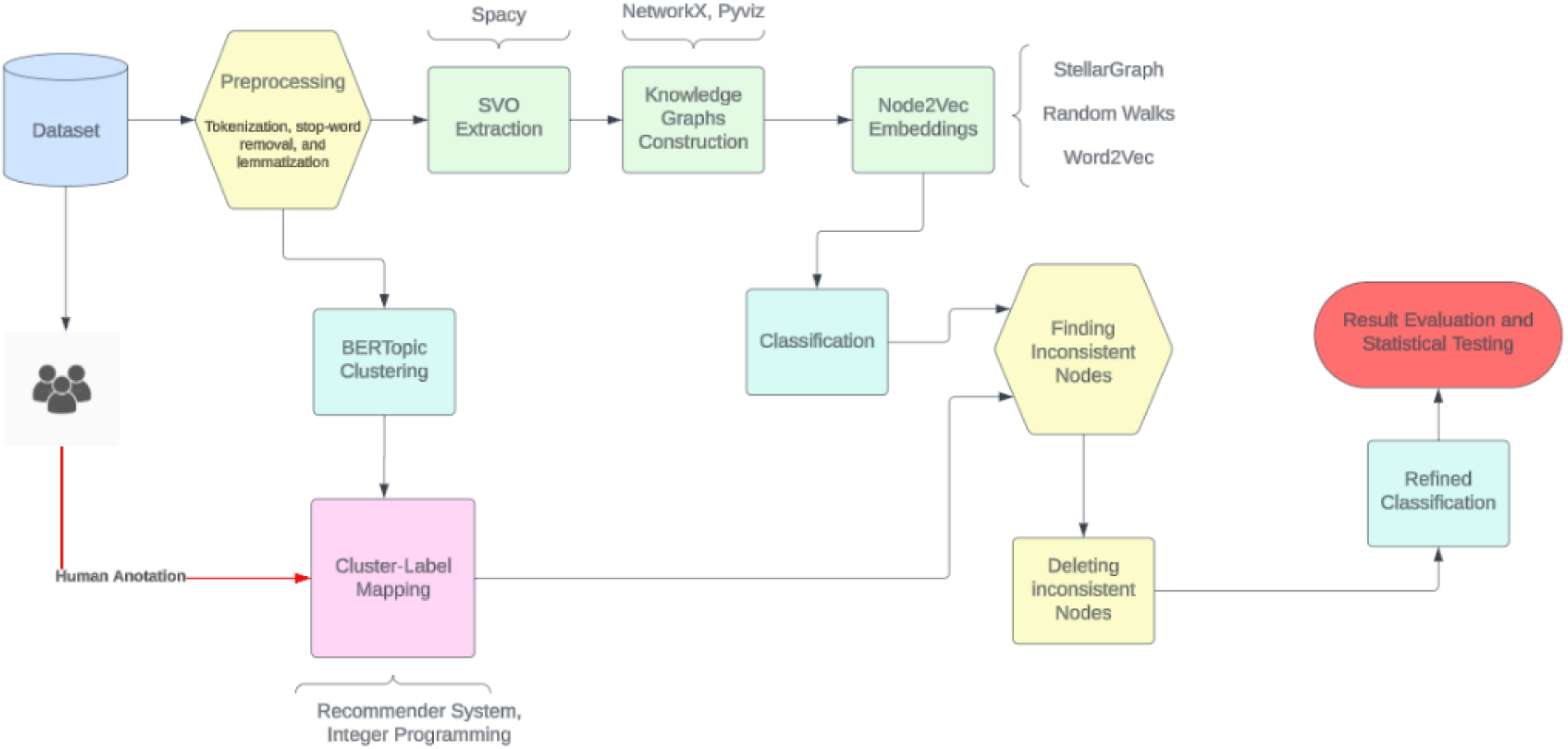
System flowchart overview.

### 2 Related Work

Several combinations of machine learning (ML) algorithms with BERT have been explored to improve classification performance.

Kassner and Schütze [2020] introduced BERT-kNN, an approach designed to enhance BERT’s performance in open-domain question answering (QA) by incorporating a k-nearest-neighbor (kNN) search component. This method significantly improves recall of facts encountered during training without requiring additional training for the BERT model. BERT-kNN excels at identifying the correct categories of responses and retrieving factually accurate answers, especially for rare facts.

Liu et al. [2020] proposed K-BERT, a language representation model that integrates knowledge graphs (KGs) into BERT to incorporate domain-specific knowledge. K-BERT mitigates the issue of knowledge noise, which can lead to deviations in meaning when excessive domain knowledge is incorporated. It achieves this through soft-position and visible matrix mechanisms, enabling effective knowledge injection without compromising the original sentence’s context.

Both of these works sequentially amalgamate their models, meaning the output of the first algorithm is passed as input to the next.

Building on this, Lin et al. [2022] introduced BertGCN, a model that combines the benefits of large-scale pretraining with transductive learning for text classification. By constructing a heterogeneous graph and representing documents as nodes using BERT embeddings, BertGCN leverages BERT’s comprehensive language understanding and graph convolution networks (GCNs) to propagate label influence, improving performance on text classification tasks. This model introduces a parallel approach by interpolating the results from GCN and BERT using a weighting factor *λ*, adding another dimension to model integration.

In a different direction, Naseem et al. [2023b] introduced two novel methodologies that integrate knowledge graph-based language models with nearest-neighbor models (kNN) and graph neural networks (GNNs). Their first approach leverages semantic and category information from neighboring instances via kNN, while the second approach uses GNNs to harness feature information from neighboring nodes in a graph. Empirical evaluations demonstrate that combining K-BERT with GNNs significantly improves relation extraction and classification tasks, particularly in the biomedical field. Additionally, they refined the GAT model by sequencing it with K-BERT to further enhance text classification performance.

While these approaches are innovative, none explicitly explore the idea of leveraging two decoupled perspectives on a single problem, a concept we seek to address in this work.

Figure 2 illustrates the architecture of several combined approaches involving BERT and different algorithms. On the left, we see BERT-kNN, where BERT feeds its output into a k-nearest-neighbor (kNN) classifier. The second block demonstrates K-BERT, where knowledge graphs (KG) are integrated into BERT to enhance its knowledge representation capabilities. In the third block, BertGCN combines BERT with Graph Convolutional Networks (GCN) through interpolation. Finally, the last block represents an integration of GAT and K-BERT, where BERT’s linguistic capabilities are combined with the graph attention network (GAT) to refine classification tasks, especially in NLP and biomedical contexts.

**Figure 2.**
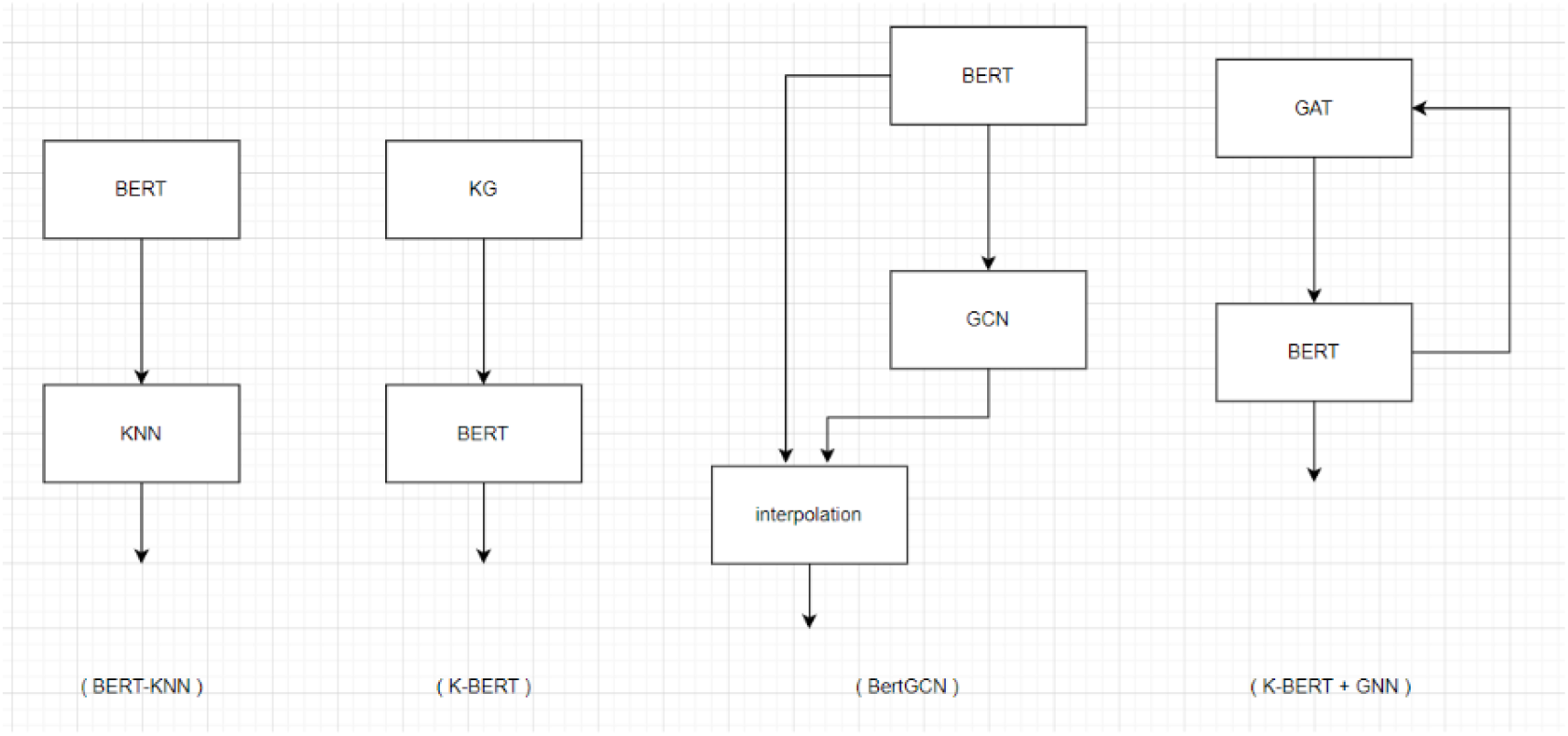
Combined BERT and Algorithmic Approaches.

## 3 Methodology

This study was conducted using a mixed methods approach which can be defined as enhancing a classifier using BERTopic insights on a text-based dataset. BERTopic generates clusters based on a large language model (LLM) [Grootendorst, 2022] which has a different insight from a traditional machine learning approach like Support Vector Machines (SVM). This combination suggests a comprehensive and potentially more insightful analysis of text data.

In the rest of this section we delve into a detailed explanation of the methodology employed with Figure 3 offering an overall visual representation of the procedural steps. Further insights into these steps are provided later in the section for a more comprehensive understanding.

**Figure 3.**
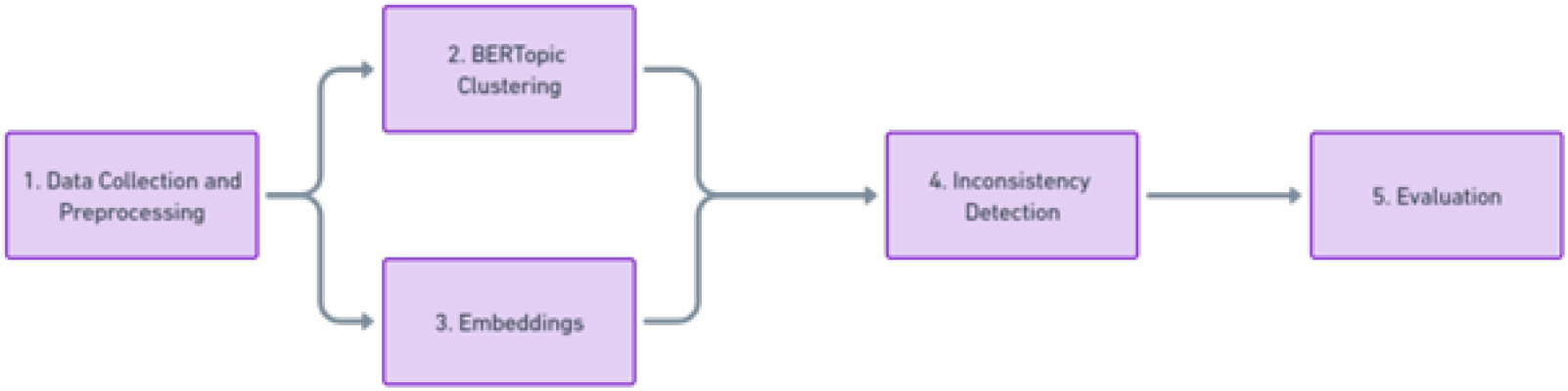
Overall workflow diagram of the methodology.

### 3.1 Data Collection and Preprocessing

The study utilized a curated dataset comprising a labeled cancer dataset^4^ with the corresponding text description from Kaggle for classification. The preprocessing phase of this study was particularly rigorous involving several key steps to ensure standardized text representations. Initially the dataset underwent tokenization, a process of breaking down text into smaller units or tokens facilitating easier analysis and processing. Following this, stop-word removal was implemented, a crucial step that involved eliminating commonly used words that offer little to no value in terms of context or meaning, thereby streamlining the dataset. Lastly, lemmatization was applied. This process involves reducing words to their base or dictionary form aiding in the consolidation of different forms of a word into a single standard form. Each of these preprocessing steps played a vital role in preparing the dataset for effective classification, ensuring a higher level of accuracy and reliability in the study’s findings.

### 3.2 BERTopic Clustering

BERTopic was used to cluster the data based on its own approach which is depicted in Figure 4. Documents are embedded into a vector space using the Sentence-BERT (SBERT) framework. This technique transforms sentences and paragraphs into dense vector representations using pre-trained language models fine-tuned for semantic similarity [Grootendorst, 2022].

**Figure 4.**
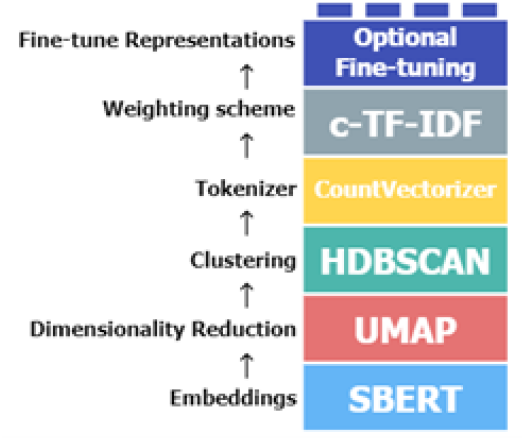
Steps to create BERTopic topic representations.^5^

For document clustering, Grootendorst [2022] addresses the challenge of high-dimensional data space where traditional distance measures become less effective. To overcome this, Grootendorst [2022] reduces the dimensionality of embeddings using UMAP (Uniform Manifold Approximation and Projection) which preserves both local and global features in lower dimensions and is adaptable to various language model dimensions [McInnes et al., 2018]. The reduced embeddings are then clustered using HDBSCAN (Hierarchical Density-Based Spatial Clustering of Applications with Noise) [McInnes and Healy, 2017]. HDBSCAN enhances the traditional DBSCAN algorithm into a hierarchical clustering model effectively differentiating between relevant clusters and outliers.

Allaoui et al. [2020] found that dimensionality reduction with UMAP improves the efficiency and accuracy of clustering algorithms like k-Means and HDBSCAN.

In this paper, when employing BERTopic there are two approaches that can be adopted. The first approach is to confine BERTopic to create as many clusters as the actual labels present in the dataset. The second approach allows BERTopic the freedom to generate any number of clusters based on its analysis of the data.

### 3.3 Embeddings

In this paper, to derive the embeddings from the text data, the steps are shown in Figure 5.

**Figure 5.**
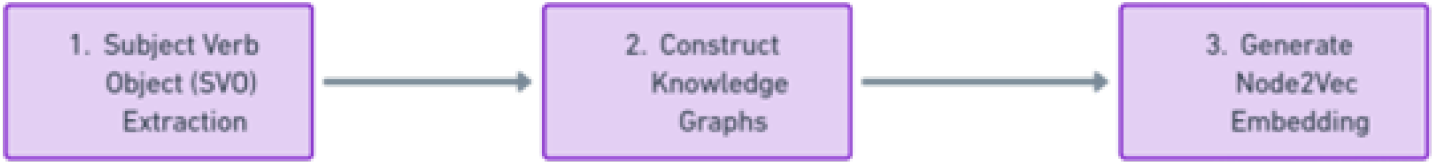
Steps to generate Embeddings.

#### 3.3.1 Subject-Verb-Object (SVO) Extraction

In order to construct a knowledge graph, we need to get the subjects, verbs, and objects of the sentences. Utilizing Spacy^6^, we can effectively identify these grammatical components. Spacy’s dependency parser allows us to parse sentences and recognize their syntactic structure making it possible to isolate the subject, verb, and object in each sentence. This is achieved by analyzing the grammatical relationships between words and identifying their respective roles.

#### 3.3.2 Knowledge Graph Construction

Once the Subject-Verb-Object (SVO) triples are extracted, they become the foundational elements for constructing the knowledge graph. In this graph, the subjects and objects are represented as nodes, while the verbs act as the edges connecting these nodes. This configuration effectively illustrates the relationships and interactions between different entities within the text. The creation of a knowledge graph from these SVO triples facilitates a visual and relational representation of the data enhancing our understanding of complex information networks within the text.

Figures 6 and 7 illustrate a sample knowledge graph that has been visualized using the NetworkX^7^ and PyViz^8^ libraries respectively.

**Figure 6.**
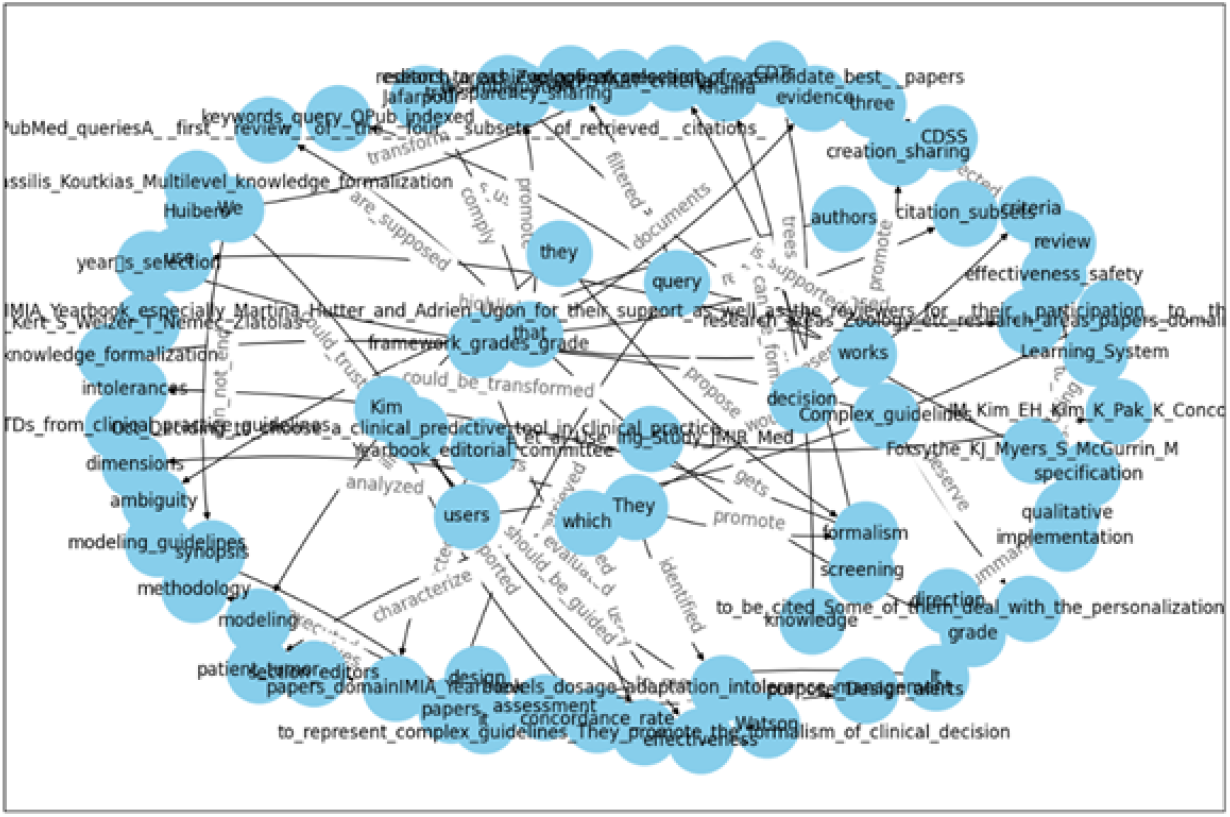
Example of a constructed knowledge graph using NetworkX.

**Figure 7.**
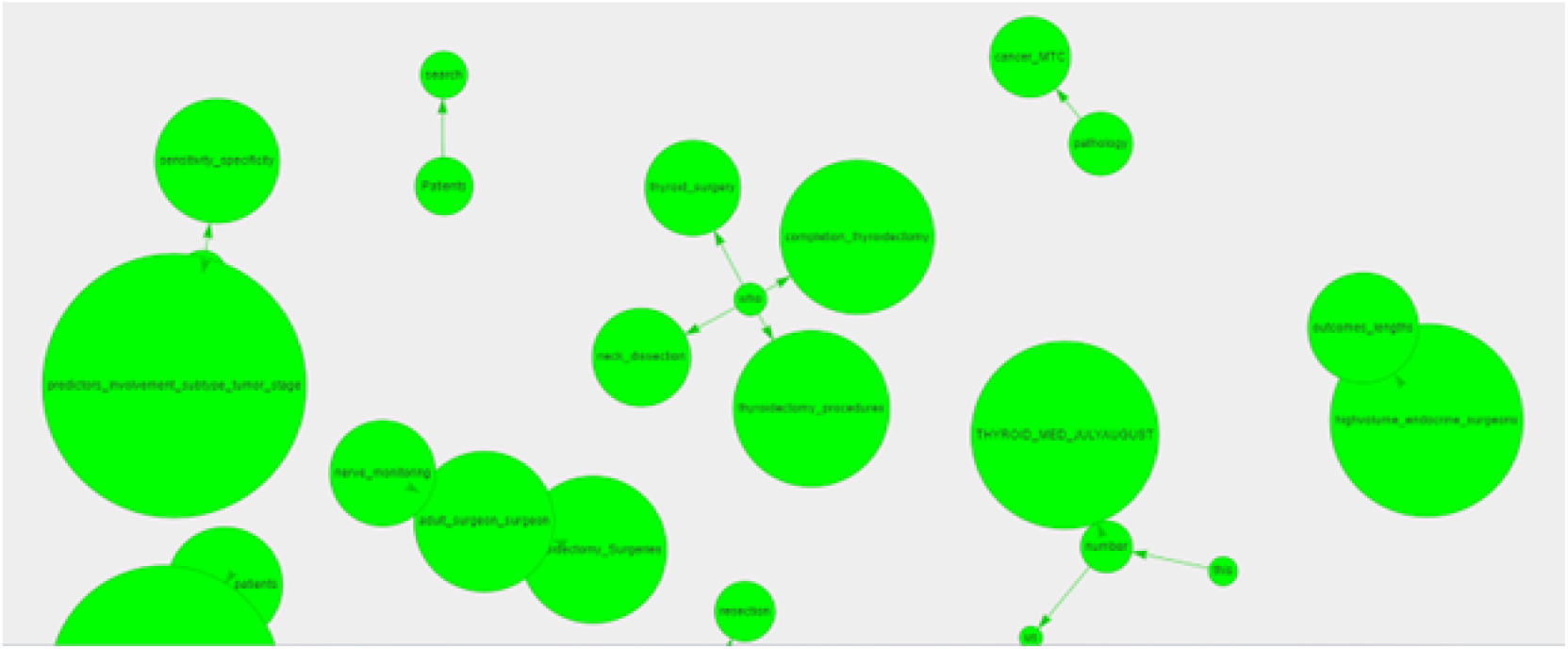
Example of a constructed knowledge graph using PyViz.

#### 3.3.3 Node2Vec Embedding Generation

In the node2vec algorithm, continuous feature representations of nodes in networks are learned by mapping nodes to a low-dimensional space. This mapping aims to maximize the likelihood of preserving network neighborhoods of nodes. The algorithm introduces a flexible notion of a node’s network neighborhood and employs a biased random walk procedure to explore these neighborhoods effectively. This flexibility allows node2vec to organize nodes based on their network roles and/or communities they belong to, adapting to various network structures and enabling more accurate representations [Grover and Leskovec, 2016].

The node2vec algorithm consists of the following steps:

1. Preprocess the graph to modify weights based on the parameters p and q, which control the random walk strategy.
2. Initialize an empty list of walks.
3. For each node in the graph, perform r random walks of length l, appending each walk to the list of walks.
4. Use Stochastic Gradient Descent (SGD) to optimize the feature representations of nodes based on the collected walks [Grover and Leskovec, 2016].

The random walks start at a node and iteratively choose the next node in the walk based on the transition probabilities that are preprocessed based on the neighborhood structure. This method efficiently samples diverse neighborhoods and return walks which are then used to learn feature representations [Grover and Leskovec, 2016].

Figure 8 depicts the random walk procedure in the algorithm.

**Figure 8.**
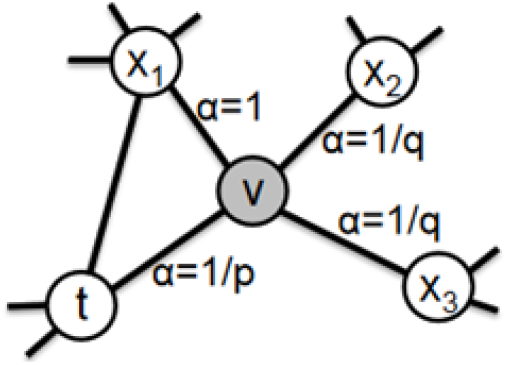
Illustration of the random walk procedure in node2vec.^9^

### 3.4 Inconsistency Detection

The next step is the innovative part of this paper which attempts to employ the results from BERTopic perspective to enhance the accuracy and F1-score of the classifier by removing the inconsistent data. Figure 9 illustrates the overall steps of the Inconsistency Detection Algorithm.

**Figure 9.**
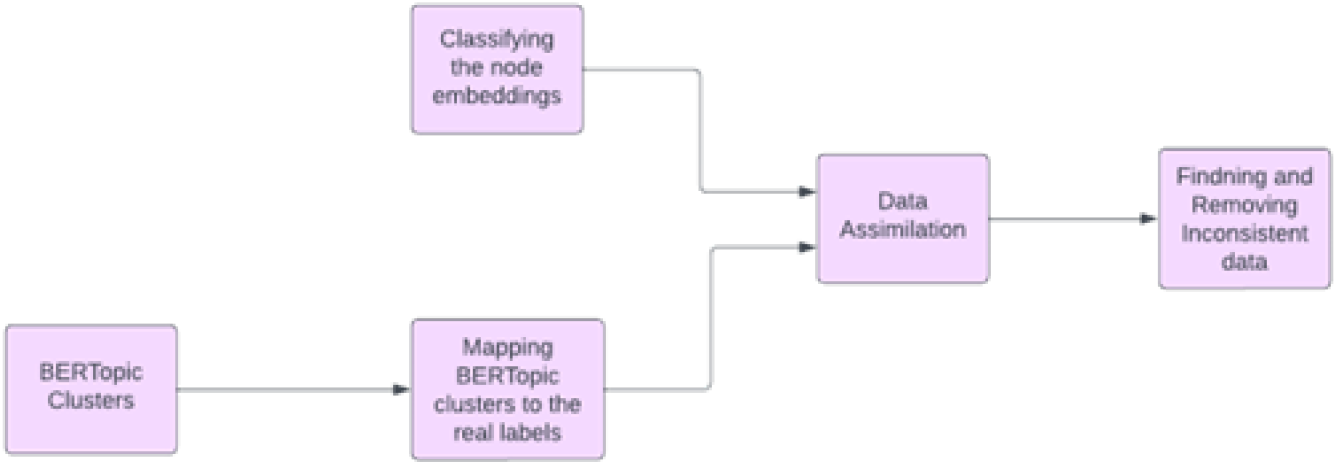
Inconsistency Detection Process.

**Figure 10.**
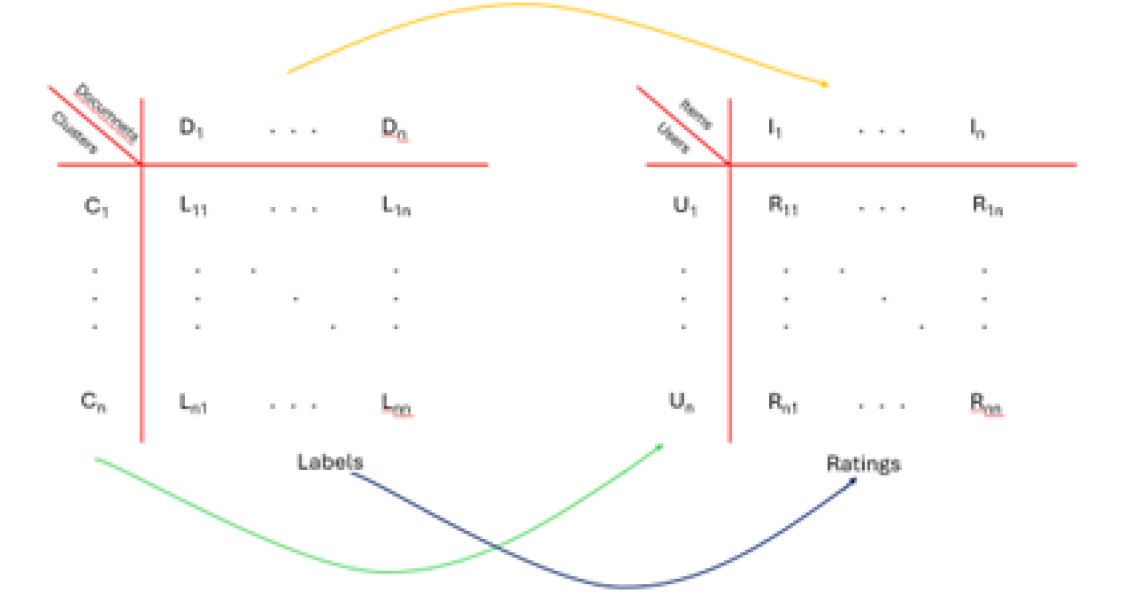
Mapping BERTopic clusters to the real labels using Recommender System.

#### 3.4.1 Mapping BERTopic Clusters to the Real Labels

The clusters generated by BERTopic are mapped to ‘real’ or ground-truth labels. This step is crucial as it aligns the clusters found in an unsupervised manner with known categories which is necessary for supervised learning tasks or to validate the clustering process.

As discussed in the “BERTopic Clustering” section, we have two distinct strategies. In the first strategy, the number of clusters generated by BERTopic equals the number of classes in our dataset. In the second strategy, BERTopic can create a diverse number of clusters. In this scenario, several clusters may correspond to a single class. Here, the main task is to identify which cluster corresponds to which class. We refer to this task as the ‘optimization process’.

Two approaches were utilized as the optimization process:

##### Class-to-Cluster Frequency Assignment

This approach involves assigning a class to the cluster where that class is most frequently observed. It’s a straightforward method that relies on the predominant presence of a class within a cluster for assignment.

- Calculate the frequency *f*_*ij*_ of class *j* in cluster *c*_*i*_:

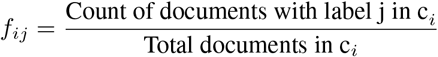
- Assign class *j* with maximum frequency to cluster *c*_*i*_:

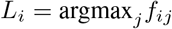

##### Recommender System Utilization

We leveraged recommender systems to determine the relationships between clusters and classes. For this, we used the Python Surprise library, a tool designed for building and analyzing recommender systems. We treated each document as an “itemID,” the clusters generated by BERTopic as “userID,” and the actual labels of the documents as “rating”. After training the model on these parameters, it predicted the associations between clusters and classes with an accuracy of 86%.

- Parameter Definitions
  * Documents: *I* = {*i*_1_, *i*_2_, …, *i*_*m*_}
  * Clusters: *U* = {*u*_1_, *u*_2_, …, *u*_*k*_}
  * Ratings (labels): *R* = {*r*_1_, *r*_2_, …, *r*_*m*_}
- Model Training
  * Train a recommender system model:

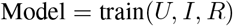
- Prediction
  * Predict cluster-to-class mapping:

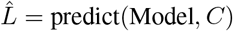

Based on the results of Tables 3 and 4 in the experimental results section, we can conclude that the recommender system results have a superior effect on the downstream classification task, improving accuracy by 34

#### 3.4.2 Classifying the Node Embeddings

Parallel to the BERTopic process, node embeddings are classified. Node embeddings are vector representations of nodes that encapsulate their properties within a graph or network. The technique used for classification in this case was Support Vector Machines (SVM).

#### 3.4.3 Data Assimilation

This phase integrates the outcomes of the BERTopic clustering with node embedding classifications. In this context, each document is treated as a distinct graph—referred to as the “graph level.” Within each graph, individual words that function as subjects, verbs, or objects are considered nodes, which constitute the “node level.”

Two assimilation strategies are presented:

- The first involves enriching the BERTopic results by transitioning from a single label per document at the graph level to multiple labels at the node level.
- The second strategy involves aggregating the labels at the node level to derive a singular label for the entire graph, determined by the most frequent node labels within that graph.

Employing either strategy sets the stage for identifying and removing inconsistent data.

#### 3.4.4 Finding and Removing Inconsistent Data

The innovative step outlined in the paper involves identifying and removing inconsistent data at the feature engineering stage, following the data assimilation process. This removal is predicated on the assumption that inconsistencies between the topic-based clusters and the network structure negatively impact the performance of classifiers. The strategic exclusion of such discordant data ensures that the classifier operates with a dataset that is more uniform and aligned, thereby potentially enhancing the precision and F1-score of the classification outcomes.

- Identify Inconsistent Data:
- Compare predicted labels *L* with actual node-level labels *L*_*n*_:

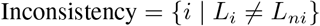
- Remove Inconsistent Data:
- Exclude inconsistent data points *I* from node embedding dataset *D*:

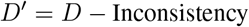

#### 3.5 Evaluation

Following the strategic exclusion of inconsistent data, the classifier is re-engaged to perform its classification tasks. Subsequently, the performance of the classifier is rigorously assessed by measuring the “Accuracy” and “F1-score”. These metrics serve as indicators of the classifier’s predictive performance.

**Accuracy** reflects the proportion of total correct predictions made by the classifier out of all predictions:

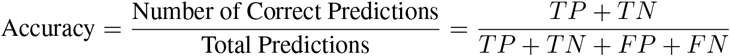

The **F1-score** provides a more nuanced measure, balancing the precision^10^ and recall^11^:

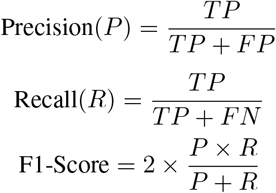

By evaluating these metrics before and after the data refinement process, we can quantify the impact of removing inconsistent data on the classifier’s effectiveness. This evaluation not only validates the methodology but also ensures that the model’s performance is aligned with the expected outcomes of the classifier post-improvement.

The conclusive phase involves conducting a **t-test**^12^ to validate that the observed improvements are statistically significant and not due to chance. This ensures that the enhanced results are not replicable by merely randomly deleting data.

- Calculate the t-statistic to validate performance improvement:

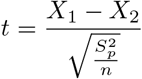

Where *X*_1_ and *X*_2_ are means of two samples, *S*_*p*_ is pooled standard deviation, and *n* is the number of samples.

## 4 Experimental Results

The strategic removal of inconsistent data, informed by BERTopic insights and subsequent reclassification, led to significant improvements in key performance metrics. To ensure that these enhancements were not simply the result of reducing data volume, we performed control experiments by randomly removing equivalent amounts of data multiple times. This control experiment aimed to determine if similar improvements could be achieved through random exclusion. We then applied a t-test to statistically validate the improvements achieved through our targeted approach, demonstrating that the improvements were specifically due to the removal of inconsistent data, rather than random data reduction.

### 4.1 BERTopic Results

Figure 11 displays the topic words derived by BERTopic for each cluster. These topic words provide insights into the clustering behavior and the content captured by the model.

**Figure 11.**
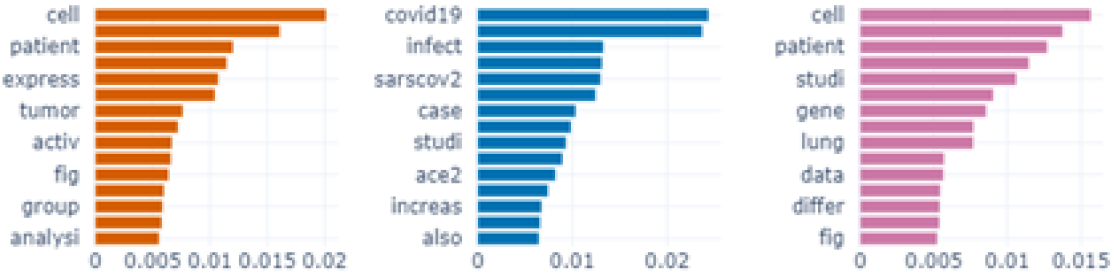
BERTopic-generated topic words for each cluster.

### 4.2 Node Embedding Visualizations

Figure 12 shows the visualization of the node embeddings, where different colors represent the true labels of the nodes. The nodes were represented using 20-dimensional vectors generated by Node2Vec, and PCA (Principal Component Analysis) was used for dimensionality reduction to enable visualization.

**Figure 12.**
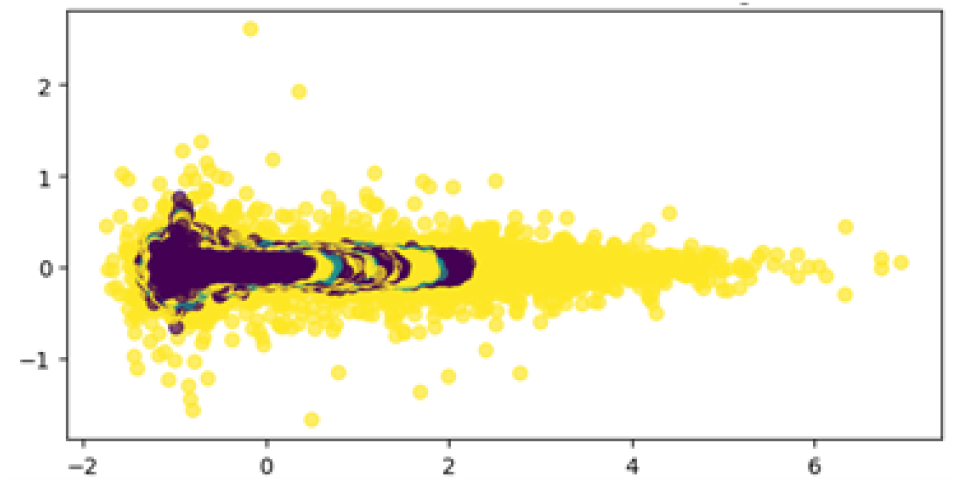
TSNE visualization of node embeddings, colored by actual labels.

### 4.3 Inconsistency Detection and Evaluation

We first classified the node embeddings using a Support Vector Machine (SVM) model. Table 2 presents the baseline accuracy and F1-score obtained before removing any inconsistent data.

**Table 1:** Node2Vec parameters considered for our case.

| Parameter | Value |
| --- | --- |
| $p$ | 0.5 |
| $q$ | 2 |
| $r$ | 5 |
| $l$ | 10 |

**Table 2:**
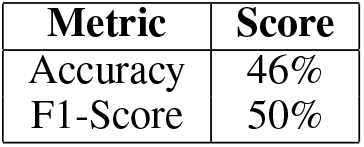
Accuracy and F1-score before removing inconsistent data.

The baseline model accuracy for this dataset was 37%, with label distribution as (2580, 2180, 2810) for each class. The SVM model’s accuracy of 46% exceeds this baseline.

Using BERTopic, we identified and flagged inconsistent data points. Figure 13 visualizes these node embeddings, with inconsistent data points clearly differentiated from the rest, illustrating their separation in the embedding space.

**Figure 13.**
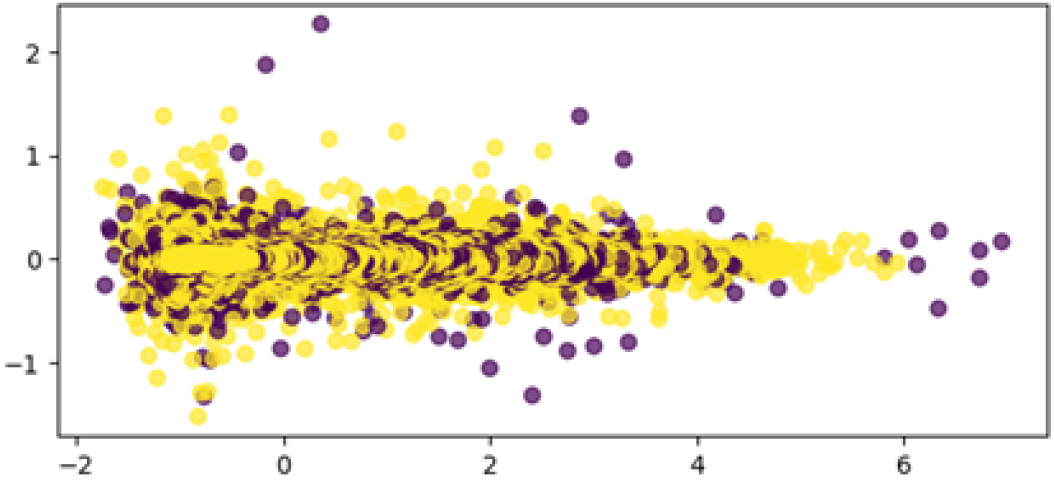
Visualization of node embeddings highlighting inconsistent data points.

After removing the inconsistent data points, we recalculated the accuracy and F1-score. As shown in Table 3, using the first optimization approach, both metrics improved by approximately 10% and 7

**Table 3:**
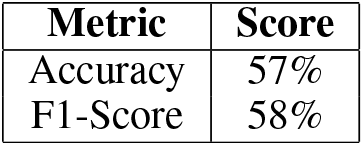
Metrics after removing inconsistent data using the first optimization approach.

**Table 4:** Metrics after removing inconsistent data using the second optimization approach.

| Metric | Score |
| --- | --- |
| Accuracy | 91% |
| F1-Score | 89% |

Figure 14 illustrates a word cloud showing the frequency of terms within the inconsistent data. Larger words in the cloud represent terms with higher frequencies, providing insight into the language characteristics of the inconsistent dataset.

**Figure 14.**
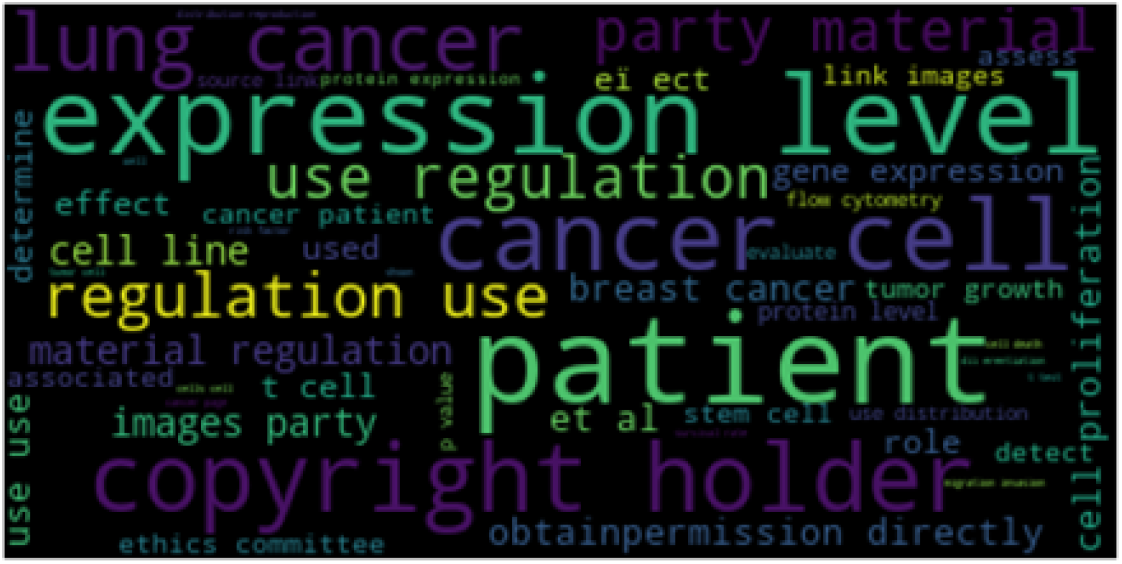
Word cloud depicting the term frequencies in inconsistent data.

Using the second optimization approach, both accuracy and F1-score improved even more significantly, with increases of approximately 18% and 16

While eliminating all inconsistencies yielded the highest improvements, a substantial portion of the data was removed in the process. To address this, we introduced a hyperparameter (*β*) to limit the removal to only *β* percent of the identified inconsistencies. With *β* = 0.7, the results shown in Table 5 were obtained, and only a smaller portion of the data was removed.

**Table 5:** Metrics after removing *β* percent of the inconsistent data.

| Metric | Score |
| --- | --- |
| Accuracy | 87% |
| F1-Score | 83% |

Lastly, we performed a t-test to assess the statistical significance of the improvements. The p-values from the t-test, shown in Table 6, demonstrate that the improvements were statistically significant.

**Table 6:**
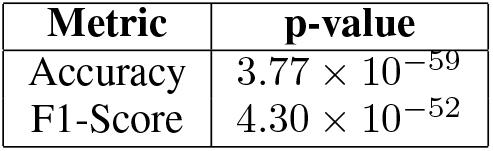
P-values from the t-test.

Since the p-values are lower than the alpha threshold of 0.05, we can confidently reject the null hypothesis. This confirms that the improvements observed are statistically significant and that the Explainable Inconsistency Algorithm was effective.

## 5 Discussion

This study set out to improve text-based cancer data classification through the combined use of BERTopic clustering and SVM classifiers, while introducing the Explainable Inconsistency Algorithm (EIA) to gain deeper insights into data inconsistencies. The integration of BERTopic with SVM significantly enhanced classification performance, with the EIA playing a critical role in detecting and explaining inconsistencies. This improved the overall reliability of the classification results.

One key innovation in this approach is the use of a recommender system for mapping clusters to labels. Employing recommender systems for this task leverages the strengths of collaborative filtering techniques—traditionally used in predicting user preferences—to enhance both the accuracy and efficiency of label assignments. This method addresses several challenges inherent in clustering and labeling tasks, offering a robust and scalable solution that adapts well to new data.

### 5.1 Handling Sparsity and High-Dimensional Data

#### Challenge

Clustering high-dimensional text data often results in sparse datasets, making it difficult to directly map clusters to labels.

#### Solution

Recommender systems excel at handling sparse data by predicting missing values and discovering latent relationships.

#### Mathematical Insight

By decomposing the association matrix *R* into lower-dimensional matrices *P* (user features) and *Q* (item features), the latent structure of the data is revealed:

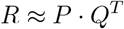

#### Equation for Matrix Factorization

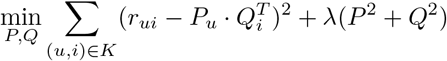

#### Result

The minimization reconstructs the rating matrix *R*, while regularizing the feature matrices *P* and *Q*, effectively managing sparsity.

### 5.2 Discovering Latent Patterns

#### Challenge

Traditional clustering methods may fail to uncover complex relationships between clusters and labels.

#### Solution

Recommender systems can discover latent factors that capture hidden relationships, improving label assignment accuracy.

#### Prediction Formula

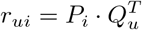

#### Explanation

The predicted rating *r*_*ui*_ reflects the likelihood of cluster *u*_*i*_ being associated with label *l*_*j*_, based on the latent factors extracted through *P* and *Q*.

### 5.3 Scalability and Efficiency

#### Challenge

Large datasets with numerous clusters and labels can be computationally demanding to process.

#### Solution

Recommender systems are designed for scalability, making them well-suited for large-scale cluster-to-label mapping.

#### Scalability Advantage

The reduced-dimensional factor matrices *P* and *Q* decrease computational complexity, making the model efficient for large datasets.

### 5.4 Flexibility and Adaptability

#### Challenge

Static clustering methods may struggle to adapt to new data or changes in data distribution.

#### Solution

Recommender systems continuously update their predictions, adapting dynamically to new clusters and labels.

#### Adaptation Formula

The model adapts as new data points (*u*_*i*_, *r*_*ui*_) are added, maintaining relevance and accuracy over time.

While BERTopic and SVM are well-established methods, this study’s unique contribution is the EIA’s ability to go beyond accuracy improvements. The EIA not only enhances performance but also facilitates the automatic detection and explanation of data inconsistencies, making it a valuable addition to the classification process.

### 5.5 Investigating Patterns using XAI

The algorithm identified several words marked as inconsistent, which may lead to confusion in model predictions, particularly in cancer-related classifications. This section explores examples of why these inconsistencies arise and how they can impact classification.

One example is the term “lung_cancer.” As shown in Figure 15, “lung_cancer” is linked to both “PTEN_expression” and “liver_cancer.” This connection can confuse the model, as PTEN mutations are commonly associated with prostate cancer, breast cancer, glioblastoma, and endometrial cancer, but less so with lung cancer.

**Figure 15.**
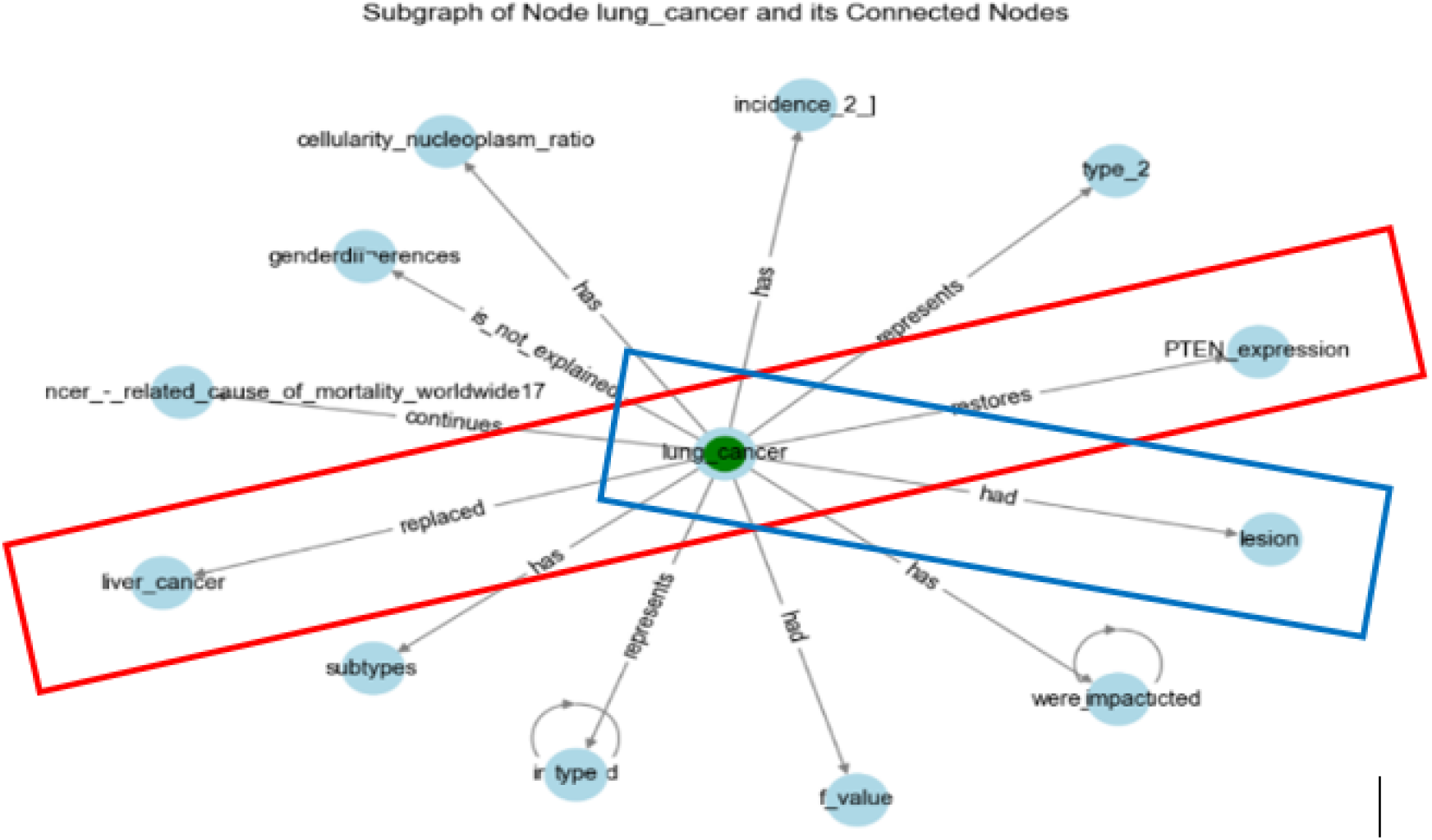
Showing the connection of “lung_cancer” to “PTEN_expression” and “liver_cancer”.

In another case, “lung_cancer” is connected to “lesion,” which is a general term for abnormal tissue. The non-specific nature of this term weakens its discriminative power, acting like a “stop word” that may lead to classification errors (Figure 16).

**Figure 16.**
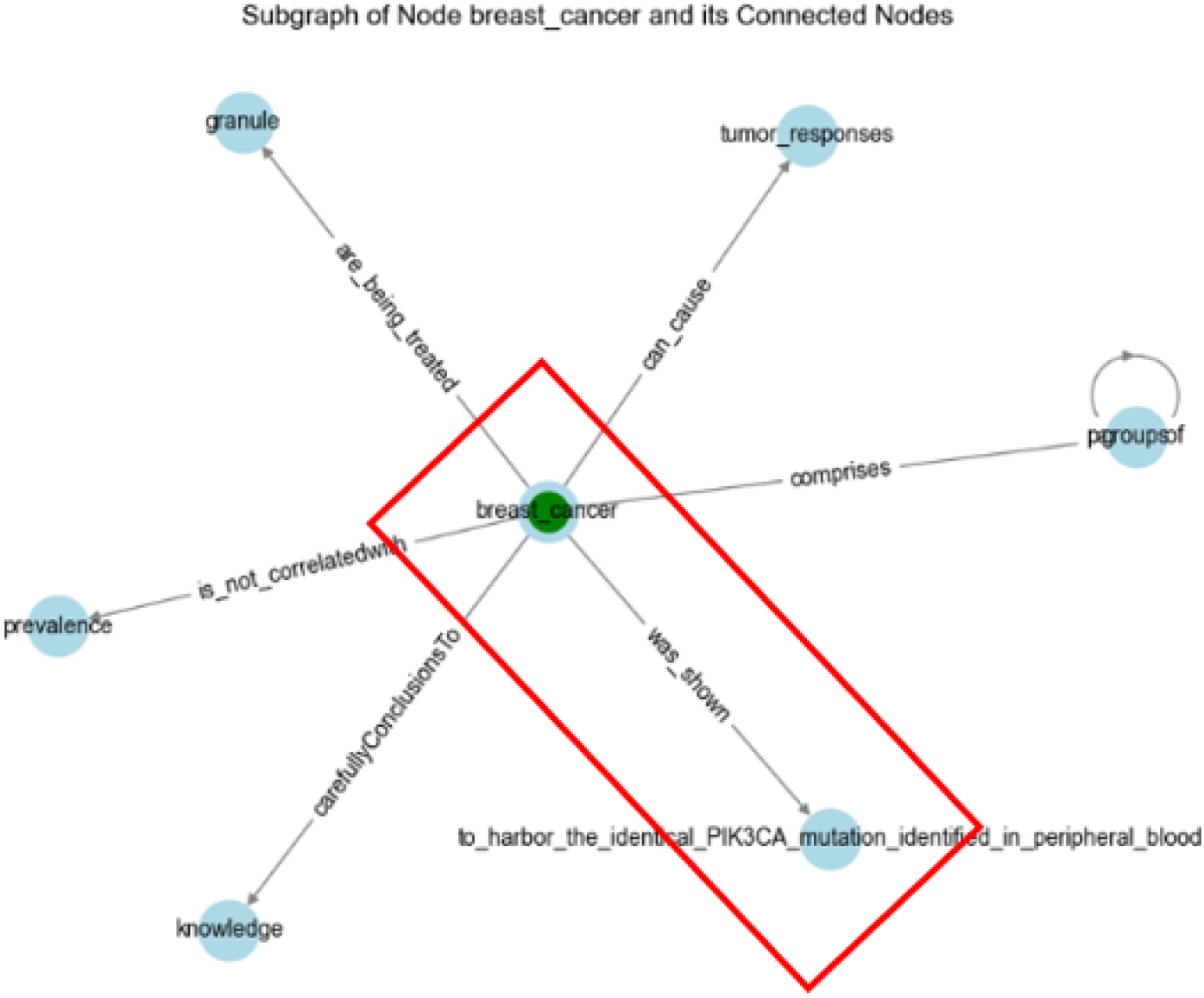
Showing connections of “lesion” in the context of cancer.

Another inconsistency is observed with “breast_cancer.” In the knowledge graph (KG), it is linked to general terms. Although some connections may seem specific, terms like “PIK3CA” are common across multiple cancers, such as lung and colon cancers, making it harder to assign this term uniquely to breast cancer.

Additionally, the term “role” appears inconsistently in the graph. The role of neutrophils in colon cancer is analogous to the role of fibrosis in lung cancer. However, “role” is a general term that connects to both cancer types, complicating the model’s ability to differentiate between them, as depicted in Figure 17.

**Figure 17.**
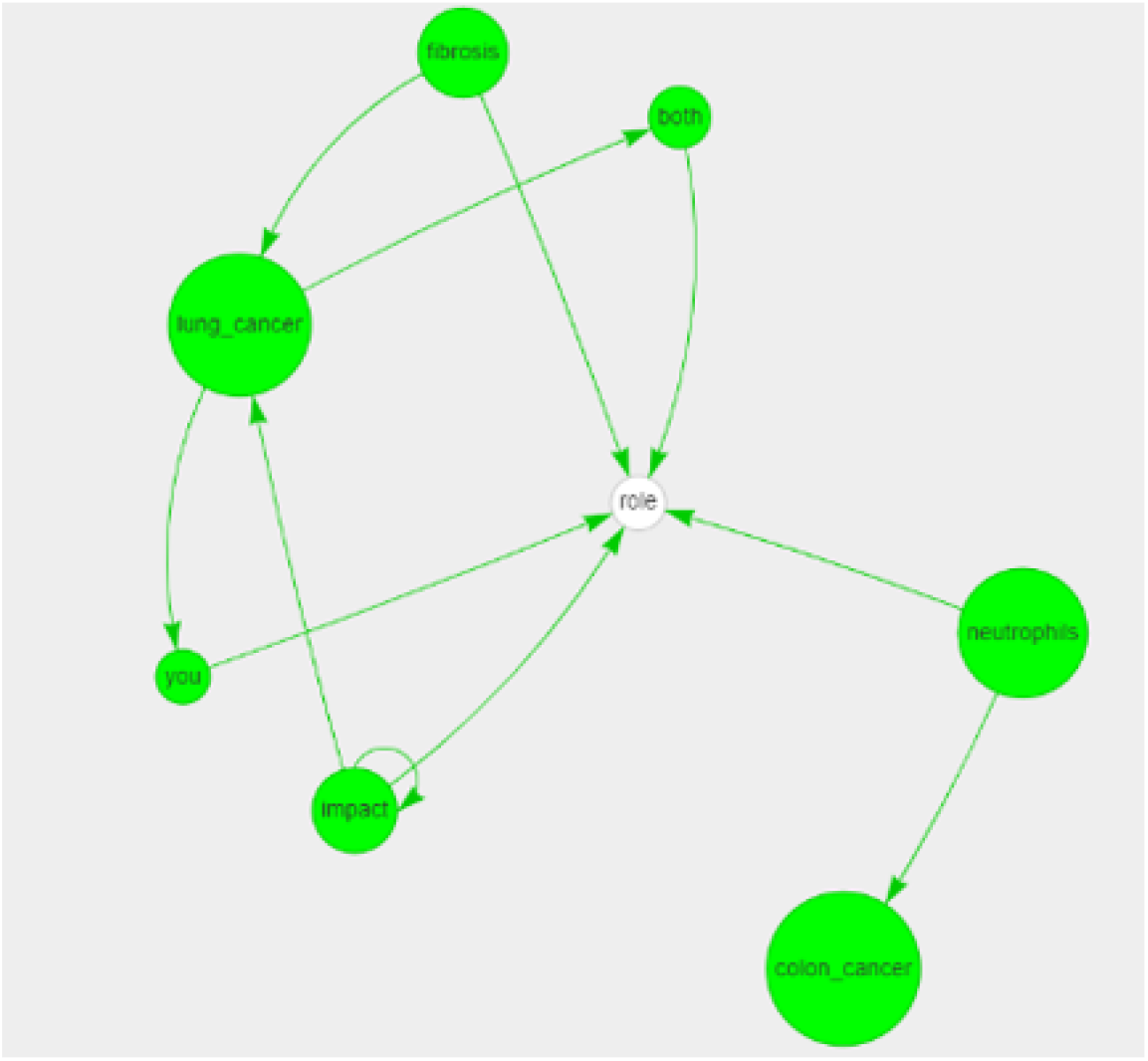
Showing the role of neutrophils and fibrosis in cancer.

Language models like BERT are better equipped to understand context compared to traditional machine learning algorithms. By analyzing node entailments in the graph, we can pinpoint sources of confusion that might mislead the model.

Furthermore, we calculated the mean degree centrality of consistent and inconsistent words. The chart below illustrates that inconsistent words exhibit much higher degree centrality:

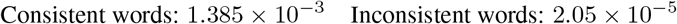

This higher degree centrality suggests that inconsistent words tend to be highly connected within the graph, often acting as hubs linking multiple terms. This high connectivity can dilute their discriminative power, leading to ambiguous predictions. In other words, the more connected a word is, the more likely it introduces noise into the model’s decision-making process, making it harder to assign clear labels.

By identifying and addressing these highly connected, yet non-discriminative nodes, the model’s performance can be significantly improved by focusing on terms that offer clear, distinctive associations with specific labels.

### 5.6 Limitations

Despite the strengths of EIA, there are limitations. The study is subject to potential dataset biases, and SVM faces challenges when dealing with complex datasets. Additionally, the use of Node2Vec, while effective for capturing relationships, leads to an increased dataset size due to the generation of numerous nodes for each document. This increase can strain computational resources and processing times, limiting scalability and efficiency in certain cases.

This study explored two distinct approaches for mapping BERTopic clusters post-Node2Vec classification. We chose the “Confining BERTopic” approach due to its superior accuracy compared to “Let BERTopic Decide.” Similarly, in the data assimilation segment, “Bringing Everything to Node Level” demonstrated higher accuracy than “Bringing Everything to Graph Level,” and was therefore selected for this research. These methodological decisions were crucial in achieving the precise and reliable results presented in this paper.

## 6. Conclusion

This research introduces an innovative approach for improving text-based cancer data classification by integrating BERTopic clustering with SVM classifiers and introducing the Explainable Inconsistency Algorithm (EIA). The proposed methodology, which leverages advanced preprocessing techniques and Node2Vec embeddings, has demonstrated significant improvements in both clustering and classification performance. More importantly, the study enabled automatic identification and removal of outliers and discordant data points using explainable methods. This integration not only enhances classification accuracy but also provides a clearer understanding of the underlying data relationships. The statistical validation through t-tests further confirms the effectiveness of the methodology, contributing substantially to the domain of text-based data analytics.

## 7 Future Work

While this study has shown promising results, several areas for future exploration could further refine and expand upon the methods developed here:

- **Integration of additional models**: Future work could explore combining other machine learning classifiers, such as Random Forests or deep learning models like Graph Neural Networks (GNNs), to evaluate whether these approaches offer further improvements when integrated with BERTopic and EIA.
- **Extension of EIA**: Expanding the Explainable Inconsistency Algorithm to different data types and domains, such as genomics or broader biomedical datasets, could enhance its applicability and effectiveness in handling data inconsistencies across various fields.
- **Optimization and scalability**: Hyperparameter tuning and optimization techniques, as well as scaling up the system for larger datasets, are key areas of focus. This could include exploring advanced parameter optimization methods and the use of parallel processing techniques for managing the computational load.
- **Real-world validation**: Deploying this methodology in a real-world clinical setting, particularly in decision-support systems, would provide valuable feedback on its performance and effectiveness in practice.

Overall, the integration of explainable machine learning techniques into cancer data classification has demonstrated substantial potential. Future work should focus on refining the approach, expanding its scope, and applying it to more diverse datasets and real-world applications.

## Data Availability

All data produced in the present work are contained in the manuscript

## Footnotes

1 https://spacy.io/

2 https://networkx.github.io/

3 https://pyviz.org/

4 https://www.kaggle.com/datasets/falgunipatel19/biomedical-text-publication-classification

5 https://maartengr.github.io/BERTopic/algorithm/algorithm.html

6 A free open-source library designed for advanced Natural Language Processing (NLP) tasks, accessible through Python. It is capable of comprehending and analyzing texts of varying sizes [JUGRAN et al., 2021]

7 A Python package for the creation and study of complex networks, offering extensive tools for network analysis and visualization.

8 A Python library for creating interactive network visualizations. It’s built on top of the popular graph library NetworkX and uses JavaScript libraries like Vis. js for rendering interactive visualizations in the browser.

9 [Grover and Leskovec, 2016]

10 The correctness of positive predictions.

11 The classifier’s ability to identify all actual positives.

12 A statistical test used to determine if there is a significant difference between the means of two groups.

## References

Thomas Hellström, Virginia Dignum, and Suna Bensch. Bias in machine learning – what is it good for? arXiv, 2020. doi:10.48550/arXiv.2004.00686.

Zengyou He, Shengchun Deng, and Xiaofei Xu. Outlier detection integrating semantic knowledge. In Xiaofeng Meng, Jianwen Su, and Yujun Wang, editors, Advances in Web-Age Information Management, Lecture Notes in Computer Science, pages 126–131. Springer, Berlin, Heidelberg, 2002. doi:10.1007/3-540-45703-8_12.

Hongzhi Wang, Mohamed Jaward Bah, and Mohamed Hammad. Progress in outlier detection techniques: A survey. IEEE Access, 7:107964–107970, 2019. doi:10.1109/ACCESS.2019.2932769.

Usman Naseem, Surendrabikram Thapa, Qi Zhang, Liang Hu, Anum Masood, and Mehwish Nasim. Reducing knowledge noise for improved semantic analysis in biomedical natural language processing applications. In Tristan Naumann, Asma Ben Abacha, Steven Bethard, Kirk Roberts, and Anna Rumshisky, editors, Proceedings of the 5th Clinical Natural Language Processing Workshop, pages 272–277, Toronto, Canada, 2023a. Association for Computational Linguistics. doi:10.18653/v1/2023.clinicalnlp-1.32.

Stefan Feldmann, Sebastian J. I. Herzig, Konstantin Kernschmidt, Thomas Wolfenstetter, Daniel Kammerl, Ahsan Qamar, Udo Lindemann, Helmut Krcmar, Christiaan J. J. Paredis, and Birgit Vogel-Heuser. Towards effective management of inconsistencies in model-based engineering of automated production systems. IFAC-PapersOnLine, 48(3):916–923, 2015. doi:10.1016/j.ifacol.2015.06.200.

Jair Cervantes, Farid Garcia-Lamont, Lisbeth Rodríguez-Mazahua, and Asdrubal Lopez. A comprehensive survey on support vector machine classification: Applications, challenges and trends. Neurocomputing, 408:189–215, 2020. doi:10.1016/j.neucom.2019.10.118.

Jiawei Wen. Efficient computing algorithm for high dimensional sparse support vector machine. arXiv, 2023. doi:10.48550/arXiv.2312.15590.

Jacob Devlin, Ming-Wei Chang, Kenton Lee, and Kristina Toutanova. Bert: Pre-training of deep bidirectional transformers for language understanding. arXiv, 2019. doi:10.48550/arXiv.1810.04805.

Santiago González-Carvajal and Eduardo C. Garrido-Merchán. Comparing bert against traditional machine learning text classification. Journal of Computational and Cognitive Engineering, April 2023. doi:10.47852/bonviewJCCE3202838.

Weiyang Liu, Yandong Wen, Zhiding Yu, and Meng Yang. Large-margin softmax loss for convolutional neural networks. arXiv, 2017. doi:10.48550/arXiv.1612.02295.

Maarten Grootendorst. Bertopic: Neural topic modeling with a class-based tf-idf procedure. arXiv, 2022. doi:10.48550/arXiv.2203.05794.

Samsir Samsir, Reagan Surbakti Saragih, Selamat Subagio, Rahmad Aditiya, and Ronal Watrianthos. Bertopic modeling of natural language processing abstracts: Thematic structure and trajectory. Jurnal Media Informatika Budidarma, 7 (3):1514–1520, 2023. doi:10.30865/mib.v7i3.6426.

Aditya Grover and Jure Leskovec. Node2vec: Scalable feature learning for networks. In Proceedings of the 22nd ACM SIGKDD International Conference on Knowledge Discovery and Data Mining, pages 855–864, New York, NY, USA, 2016. Association for Computing Machinery. doi:10.1145/2939672.2939754.

Enrico Palumbo, Giuseppe Rizzo, Raphaël Troncy, Elena Baralis, Michele Osella, and Enrico Ferro. Knowledge graph embeddings with node2vec for item recommendation. In Aldo Gangemi, Anna Lisa Gentile, Andrea Giovanni Nuzzolese, Sebastian Rudolph, Maria Maleshkova, Heiko Paulheim, Jeff Z. Pan, and Mehwish Alam, editors, The Semantic Web: ESWC 2018 Satellite Events, Lecture Notes in Computer Science, pages 117–120. Springer International Publishing, Cham, 2018. doi:10.1007/978-3-319-98192-5_22.

YueQun Wang, LiYan Dong, XiaoQuan Jiang, XinTao Ma, YongLi Li, and Hao Zhang. Kg2vec: A node2vec-based vectorization model for knowledge graph. PLOS ONE, 16(3):e0248552, 2021. doi:10.1371/journal.pone.0248552.

Nora Kassner and Hinrich Schütze. BERT-kNN: Adding a kNN search component to pretrained language models for better QA. In Trevor Cohn, Yulan He, and Yang Liu, editors, Findings of the Association for Computational Linguistics: EMNLP 2020, pages 3424–3430, Online, November 2020. Association for Computational Linguistics. doi:10.18653/v1/2020.findings-emnlp.307. URL https://aclanthology.org/2020.findings-emnlp.307.

Weijie Liu, Peng Zhou, Zhe Zhao, Zhiruo Wang, Qi Ju, Haotang Deng, and Ping Wang. K-bert: Enabling language representation with knowledge graph. Proceedings of the AAAI Conference on Artificial Intelligence, 34(03):2901–2908, Apr. 2020. doi:10.1609/aaai.v34i03.5681. URL https://ojs.aaai.org/index.php/AAAI/article/view/5681.

Yuxiao Lin, Yuxian Meng, Xiaofei Sun, Qinghong Han, Kun Kuang, Jiwei Li, and Fei Wu. Bertgcn: Transductive text classification by combining gcn and bert, 2022. URL https://arxiv.org/abs/2105.05727.

Usman Naseem, Surendrabikram Thapa, Qi Zhang, Liang Hu, Anum Masood, and Mehwish Nasim. Reducing knowledge noise for improved semantic analysis in biomedical natural language processing applications. In Tristan Naumann, Asma Ben Abacha, Steven Bethard, Kirk Roberts, and Anna Rumshisky, editors, Proceedings of the 5th Clinical Natural Language Processing Workshop, pages 272–277, Toronto, Canada, July 2023b. Association for Computational Linguistics. doi:10.18653/v1/2023.clinicalnlp-1.32. URL https://aclanthology.org/2023.clinicalnlp-1.32.

Leland McInnes, John Healy, Nathaniel Saul, and Lukas Großberger. Umap: Uniform manifold approximation and projection. Journal of Open Source Software, 3(29):861, 2018. doi:10.21105/joss.00861. URL 10.21105/joss.00861.

Leland McInnes and John Healy. Accelerated hierarchical density based clustering. In 2017 IEEE International Conference on Data Mining Workshops (ICDMW). IEEE, November 2017. doi:10.1109/icdmw.2017.12. URL 10.1109/ICDMW.2017.12.

Mebarka Allaoui, Mohammed Lamine Kherfi, and Abdelhakim Cheriet. Considerably improving clustering algorithms using umap dimensionality reduction technique: A comparative study. In Abderrahim El Moataz, Driss Mammass, Alamin Mansouri, and Fathallah Nouboud, editors, Image and Signal Processing, pages 317–325, Cham, 2020. Springer International Publishing. ISBN 978-3-030-51935-3.

Swaranjali Jugran, Ashish Kumar, Bhupendra Singh Tyagi, and Vivek Anand. Extractive auto-matic text summarization using spacy in python nlp. In 2021 International Conference on Advance Computing and Innovative Technologies in Engineering (ICACITE), pages 582–585, 2021. doi:10.1109/ICACITE51222.2021.9404712.

